# COVID-19 vaccination and survival of cancer patients in Northwestern Russia: a historical registry-based cohort study

**DOI:** 10.64898/2026.10.12.26365129

**Authors:** Andrey A. Dyachenko, Maxim A. Bogdanov, Dmitry V. Bogdanov, Anna A. Meldo, Valeria Yu. Chernina, Victor A. Gombolevskiy, M. Grjibovski, Mikhail Yu. Valkov

## Abstract

**Background:** Cancer patients face a significantly elevated risk of severe COVID-19 outcomes. Although vaccination is a cornerstone of prevention, data on vaccination coverage and association with survival among Russian cancer patients remain scarce. We assessed COVID-19 vaccination coverage and its association with overall survival in a population-based cohort of cancer patients in Northwest Russia.

**Methods:** This retrospective registry-based cohort study integrated data from the Population-Based Cancer Registry (PBCR) of the Arkhangelsk Region and the Nenets Autonomous Okrug, regional vaccination records, and an artificial intelligence (AI)-based analysis of chest computed tomography (CT) scans. The cohort consisted of 1,144 cancer patients who underwent CT between August 2020 and April 2021. Vaccination status was ascertained via data linkage. Overall survival (OS) was compared between vaccinated and unvaccinated patients using Kaplan–Meier analysis and multivariable Cox proportional hazards regression, adjusted for demographic, clinical, and AI-derived CT biomarkers. To mitigate immortal time bias, the primary analysis used a landmark approach restricted to patients alive on March 1, 2021 (N = 1,136). A sensitivity analysis used a November 1, 2021 landmark (N = 894). Vaccination status was defined as receipt of at least one dose before the landmark date.

**Results:** Only 6.6% (n=76) of patients received at least one dose of a COVID-19 vaccine (Sputnik V or Sputnik Light). Vaccinated patients were more likely to have earlier-stage cancer and were less likely to be receiving active treatment. In the primary landmark analysis, vaccination was associated with a significantly higher 3-year OS (80.3% vs. 45.7%, p < 0.001). In multivariable analysis, vaccination was independently associated with a 60% lower hazard of all-cause mortality (adjusted HR 0.40, 95% CI 0.25–0.64). AI-detected COVID-19 pneumonia on baseline CT was also an independent predictor of poorer survival (adjusted HR 1.32, 95% CI 1.09–1.59). In the sensitivity analysis (November 1, 2021 landmark), the adjusted HR for vaccination was 0.56 (95% CI 0.34–0.92), showing a consistent but attenuated association.

**Conclusion:** This study reveals critically low COVID-19 vaccination coverage among cancer patients in Northwest Russia. Nevertheless, vaccination was strongly associated with a substantial survival benefit, underscoring its importance as a potentially lifesaving intervention for this vulnerable population.

## Introduction

The global impact of COVID-19 has been profound and multifaceted, extending far beyond the immediate health crisis to trigger economic, social, and geopolitical shocks. According to data from WHO COVID-19 dashboard, as of Aug 2025, there were 778 million officially reported cases and more than 7 million deaths from COVID-19 [1]. In Russia, excess mortality associated with the pandemic in 2021 was estimated as 41.2% overall [2].

Patients with cancer are at a significantly higher risk of severe illness, hospitalization, and death from COVID-19 than the general population. This increased risk is driven by comorbidities, poor performance status, the presence of hematological malignancies, and receipt of immunosuppressive cancer therapies [3; 4]. During the pandemic, preventing infection and severe COVID-19 was critical, and vaccination remained the most effective strategy. Global efforts led to the rapid development of several highly effective vaccines. Although mass vaccination has markedly reduced the incidence of severe COVID-19 worldwide, cancer patients—who are at high risk—were appropriately prioritized for vaccination in most countries [*5; 6*], including Russia [*7*]. Although vaccinated patients with cancer were more likely to develop breakthrough SARS-CoV-2 infections with severe outcomes [8], vaccination was still associated with improved survival among patients with cancer compared with no vaccination [9].

Global COVID-19 vaccination coverage of among patients with cancer has varied widely. Evidence from Western Europe indicates a significant increase in uptake over time. In the OnCovid hospital-based registry, Pinato et al. found that although only 16.6% of patients with cancer were fully vaccinated during the Alpha/Delta phase of the pandemic, this figure rose to 33.3% among those diagnosed with COVID-19 during the later Omicron phase, reflecting improving but still suboptimal coverage [10]. Globally, a meta-analysis of 29 studies by Prabani et al., reported a pooled acceptance for at least one vaccine dose of 59.0% (95% confidence interval: 52–67%, I²: 99%) among patients with cancer [11]. In contrast to these figures, specific and reliable data on vaccination coverage among Russian cancer patients remains lacking in the international peer-reviewed literature.

Radiologic signs suggestive of COVID-19 pneumonia that are discovered incidentally on CT scans in cancer patients without typical clinical symptoms (i.e. asymptomatic or pre-symptomatic presentation) are associated with a significantly higher risk of disease progression and mortality [12]. In our earlier population-based cohort study, which used artificial intelligence (AI)-powered data processing for including patients with malignant neoplasms who underwent chest CT in Arkhangelsk, Northwestern Russia, the presence of CT signs of COVID-19 was associated with a 31% increased risk of all-cause mortality [13]. We therefore integrated this cohort with regional COVID-19 vaccination records for subsequent analysis of outcomes.

Primary Objective: To assess COVID-19 vaccination coverage and its association with overall survival among cancer patients in a population-based cohort.

Secondary Objectives:

- To characterize the demographic and clinical profiles of vaccinated versus unvaccinated patients with cancer.
- To compare AI-derived CT findings and imaging biomarker categories between vaccination groups and to assess their associations with overall survival.

## Methods

This historical registry-based cohort study is a part of the ARILUS project [14], which investigates the use of multipurpose AI in CT to reduce non-cancer-related mortality in oncology patients. The study was approved by the Local Ethics Committee of the Northern State Medical University (Protocol No. 07/10-238, 2023).

### Data Collection

All chest CT series performed between August 1, 2020, and April 30, 2021, corresponding to the COVID-19 pandemic period, were extracted from the central medical image archive of the Arkhangelsk Clinical Oncology Dispensary (ACOD). In Russia, an oncology dispensary is a comprehensive medical institution dedicated to the prevention, diagnosis, and treatment of cancer, including long-term follow-up care for patients who have completed treatment.

During the pandemic period, chest CT was prescribed for two main indications: (1) routine diagnostic clarification and assessment of the prevalence of malignant neoplasms (MNs), and (2) evaluation for possible viral pneumonia if a patient developed COVID-19 symptoms during hospitalization. At the time of hospitalization, patients were required to be asymptomatic and have a negative nasopharyngeal swab test for COVID-19 virus. After exclusion of CTs performed for non-malignant conditions, a total of 7,620 CT studies were identified in cancer patients during this period.

For patients with multiple eligible CT scans, only the first scan during the study period was included. Selection of images valid for processing was based on the limitations of the AI algorithm: cases of lung atelectasis were excluded based on radiologists’ description protocols, and CT series with intravenous contrast were excluded based on analysis of the DICOM tag (Protocol Name Tag 0018,1030). After de-identification, all valid CT series were sent for AI analysis to IRA Labs LLC (Moscow, Russia). At this stage, cases with severe motion artifacts, slice thickness greater than 1.5 mm, and incomplete lung scanning zone were excluded. The total number of patients with valid CT studies was 1,542.

After AI processing, the results were sent to ACOD via a secure channel using the insurance number of an individual personal account (SNILS, Rus: СНИЛС – страховой номер индивидуального лицевого счёта) and merged with the database of the Population-Based Cancer Registry (PBCR) of the Arkhangelsk Region and the Nenets Autonomous Okrug, extracted on April 15, 2025. The general PBCR database contained information on 146,742 patients registered with a diagnosis of invasive MN as of the extraction date. Extracting data from the PBCR was necessary to establish the causes of death for patients for whom SNILS data were available. Among the patients with valid CT studies, 395 patients had MNs, but their SNILS data were unavailable, making outcome assessment impossible; these patients were excluded. We also excluded three additional records because their CT scans were conducted before the pandemic in Arkhangelsk (Figure 1).

**Figure 1.**
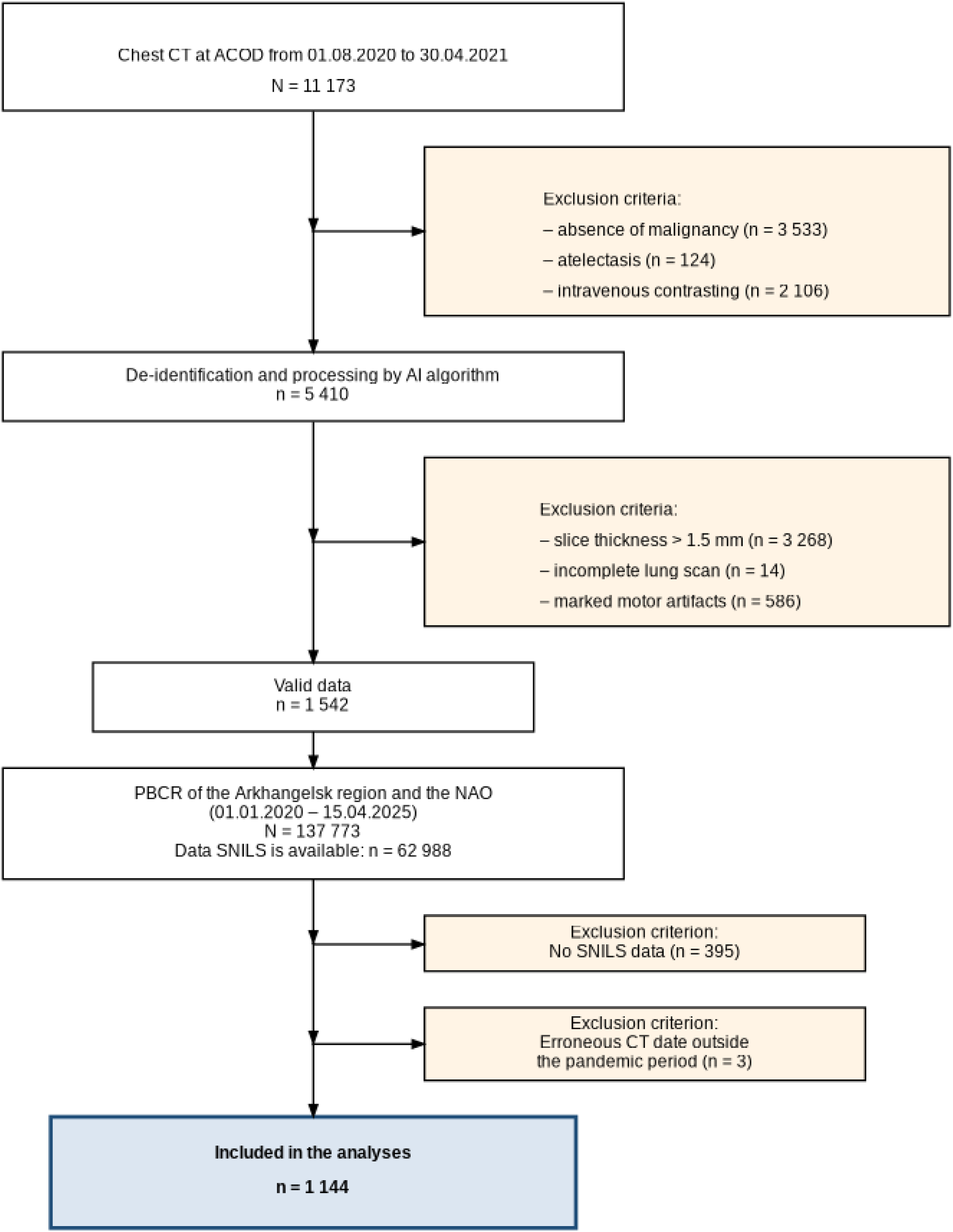
Flowchart of the study. Notes: CT – computed tomography; ACOD – Arkhangelsk Clinical Oncological Dispensary, AI – artificial intelligence, PBCR – population–based cancer registry, NAO – Nenets Autonomous Okrug, SNILS – (Rus: СНИЛС, страховой номер индивидуального лицевого счета) insurance number of individual personal account.

### Vaccination Data Collection

Vaccination data were obtained from the regional unified health information system (UHIP) of the Arkhangelsk Oblast and the Nenets Autonomous Okrug. Data extraction was performed on April 15, 2025, and included all records of administered COVID-19 vaccines available in the Russian Federation (Gam-COVID-Vac (Sputnik V, Sputnik Light), CoviVac, EpiVacCorona) for the period from January 2021 to December 2023.

Data linkage with the study cohort was performed using SNILS identifier. For patients with multiple vaccine records, the date of the first administered dose was considered the beginning of the vaccination course.

### Multipurpose AI Algorithm

To detect and assess infiltrative changes in the lungs (both qualitative and quantitative, including the percentage of involvement for each lung separately) characteristic of COVID-19-associated viral pneumonia (ICD-10 code U07.1), AI-based software – “Intelligent Radiology Assistants” (developer: IRA Labs LLC, Moscow, Russia) – was used. This medical device complies with technical specifications TU 58.29.32-001-44270315-2021 and holds the Roszdravnadzor registration certificate No. RZN 2024/22895. This algorithm was used during the pandemic in medical institutions providing primary healthcare to the adult population in Moscow for the automated analysis of chest CT studies, which helped standardize diagnosis and objectively assess the extent of lung damage [15]. Each AI result included two CT series: images and texts in DICOM SR format. Prior to this, the AI was evaluated on independent data using closed datasets as part of the Moscow Experiment [16]. The algorithm’s accuracy was 0.98, sensitivity 0.94, and specificity 0.94.

The algorithm is multipurpose, enabling simultaneous detection of several different pathologies on chest CT scans, including pulmonary emphysema, measurement of aortic and pulmonary trunk diameter, coronary artery calcification, epicardial fat volume, and CT attenuation of vertebral bodies within the scanned field of view. Figure 2 shows an example of AI processing.

**Fig. 2.**
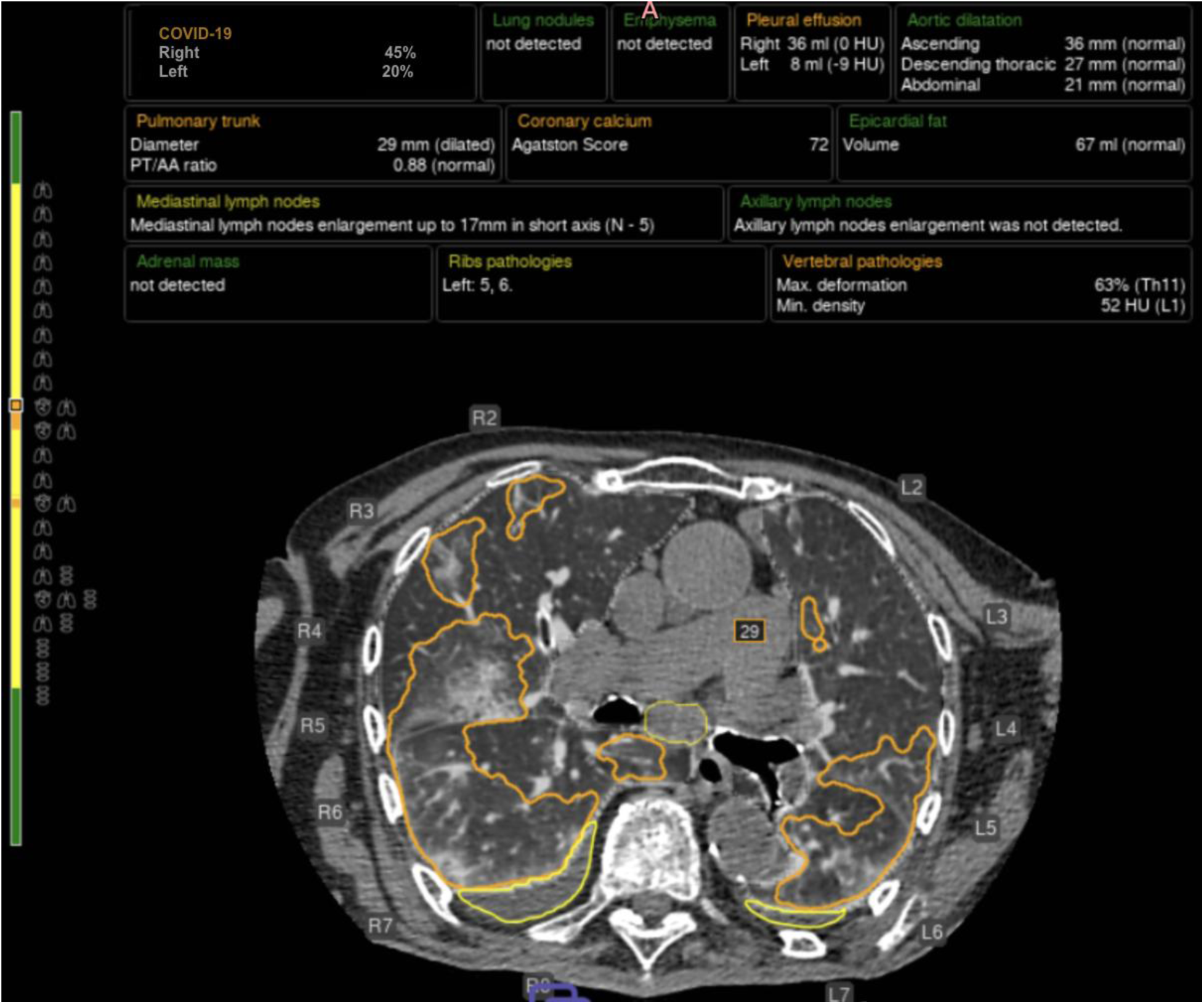
Processing of chest computed tomography images using multi-target artificial intelligence technology. Lung tissue lesions of COVID-19-associated pneumonia are presented as percentages for each lung. Fluid in the pleural cavity (yellow), its volume and density, vessel diameters (including orange for the enlarged pulmonary trunk), and outlines of enlarged mediastinal lymph nodes (pale yellow) are also shown.

The COVID-IRA module underwent independent external validation. Its performance metrics, the validation dataset, and the reference standard are described in Appendix 1.

### Variables

The unified database integrated SNILS-linked records from the PBCR, outputs from an AI algorithm, and vaccination data. It contained the following variables:

- *Demographics*: Unique patient identifier (SNILS), sex, date of birth, and type of residence (urban/rural).
- *Cancer Characteristics*: ICD-10 code, date of diagnosis, ICD-O-3.1 morphological code, clinical stage, full TNM-8 classification (including individual T, N, M categories and the overall stage group), and a variable indicating active treatment (any treatment administered within the last two months before CT scan).
- *Chest CT AI Analysis Results*: CT scan date, diameters (mm) of the ascending aorta, descending aorta, abdominal aorta, and pulmonary trunk, coronary artery calcium (CAC) Agatston score, epicardial adipose tissue volume (mL), percentage of lung tissue affected by emphysema, and the minimum mean vertebral density for assessing osteoporosis/osteopenia.
- *Vaccination Details*: Date of administration, vaccine name, batch number, and administering medical facility code.
- *Survival Data*: Vital status (alive, deceased, lost to follow-up), date of death, and cause of death.

### Statistical analysis

A comparison was performed between two cohorts of cancer patients: those vaccinated against COVID-19 during the pandemic and those unvaccinated. Overall survival (OS), defined as the time *from the CT scan* date to death from any cause, was used as the primary outcome measure. Patients who remained alive were censored on February 15, 2025 -a date two months prior to the most recent update of patient status data. Information on patients in the PBCR of the Arkhangelsk Oblast and Nenets Autonomous Okrug is updated monthly by cross-referencing registry data with the regional mortality database.

COVID-19 vaccination became widely available to the general population in Russia starting in early January 2021. To mitigate immortal time bias — whereby patients who died early could not have been vaccinated — the primary survival analysis used a landmark approach restricted to patients who were alive and under observation as of March 1, 2021. This landmark date also excluded terminally ill cancer patients with a life expectancy of less than one-two month who would have been ineligible for vaccination per vaccine labeling. Additionally, a sensitivity analysis was performed using a cut-off date of November 1, 2021, which corresponds to two months after the issuance of a federal directive prioritizing cancer patients for COVID-19 vaccination, allowing time for the directive to be implemented in clinical practice.

Categorical variables were analyzed using Pearson’s chi-squared test. Survival analysis was performed using the Kaplan-Meier method. Differences in survival rates were compared using the log-rank test. Univariate comparisons of survival functions were conducted using the Mantel–Cox log-rank test. Multivariable Cox proportional hazards regression was used to calculate hazard ratios with 95% confidence intervals (95% CI) adjusted for potential confounders [17]. A p-value of <0.05 was considered statistically significant. Statistical analysis was performed using Stata v.18 software (Stata Corp., TX, USA).

## Results

A total of 76 (6.6%) patients in the cohort received at least one dose of a COVID-19 vaccine. The vaccinated patients received one of two Russian adenovirus-vectored vaccines: the two-dose Sputnik V (Gam-COVID-Vac, n=48) or the single-dose Sputnik Light (Gam-COVID-Vac-Light, n=28). No patients received CoviVac or EpiVacCorona. The distribution of vaccinated and unvaccinated patients across key demographic and clinical characteristics, including CT-derived bioindicators identified by an AI algorithm, is presented in Table 1.

**Table 1.**
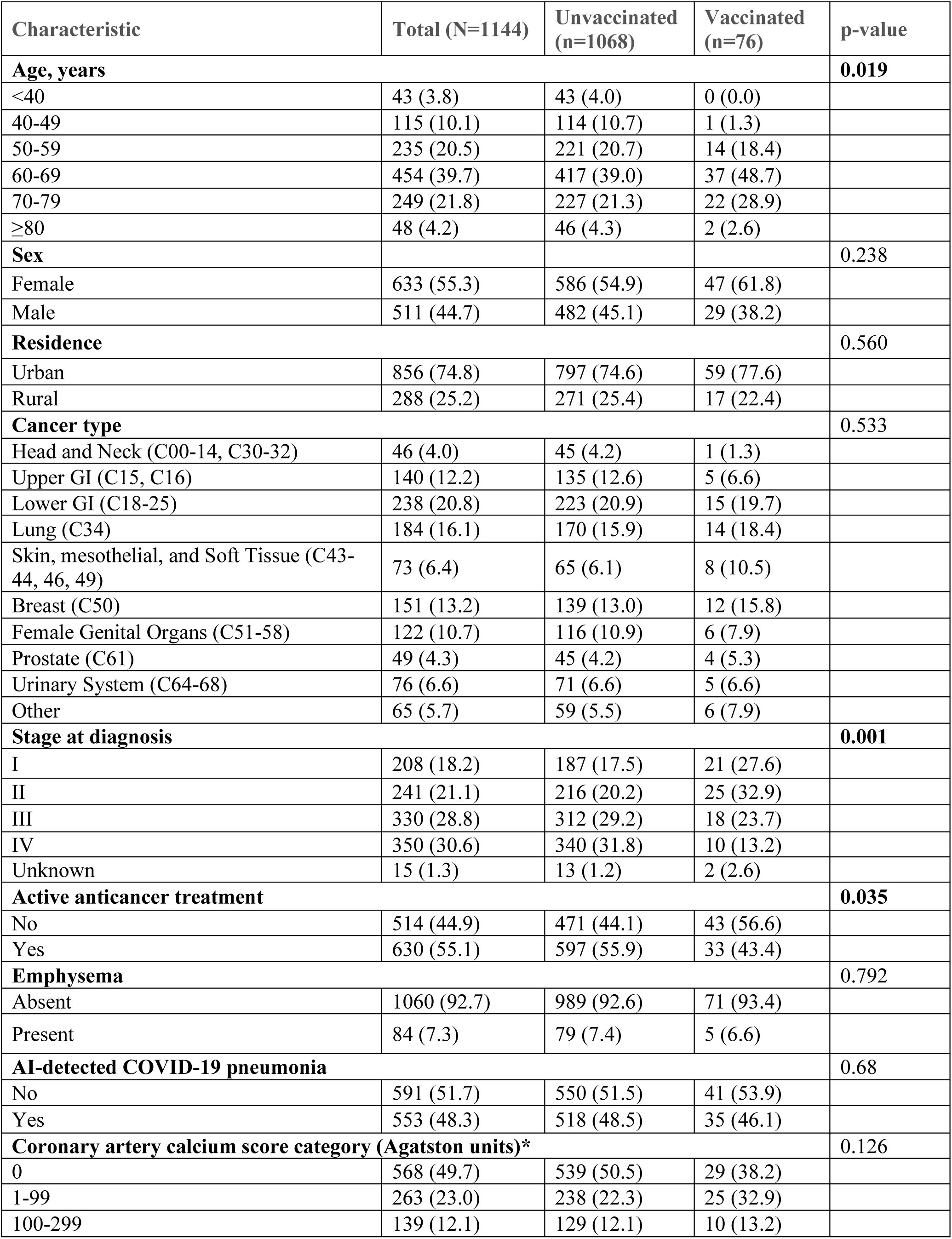
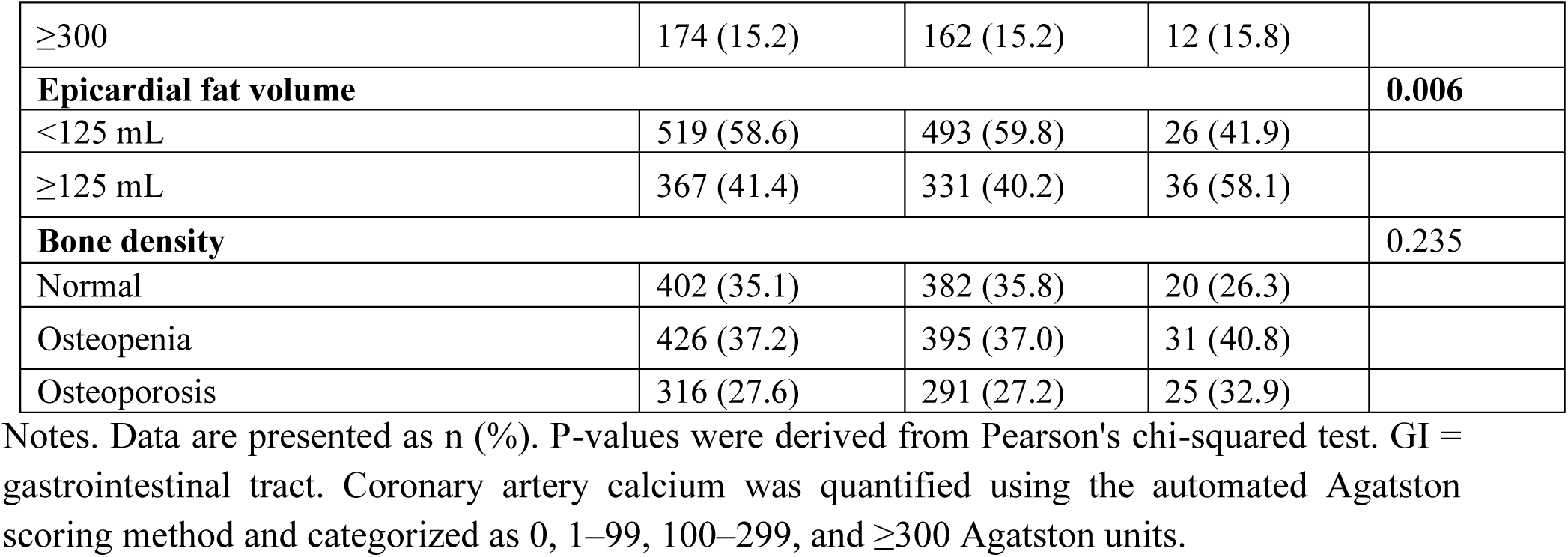
Baseline Demographic and Clinical Characteristics of Cancer Patients by COVID-19 Vaccination Status

| Characteristic | Total (N=1144) | Unvaccinated (n=1068) | Vaccinated (n=76) | p-value |
| --- | --- | --- | --- | --- |
| **Age, years** |  |  |  | **0.019** |
| <40 | 43 (3.8) | 43 (4.0) | 0 (0.0) |  |
| 40-49 | 115 (10.1) | 114 (10.7) | 1 (1.3) |  |
| 50-59 | 235 (20.5) | 221 (20.7) | 14 (18.4) |  |
| 60-69 | 454 (39.7) | 417 (39.0) | 37 (48.7) |  |
| 70-79 | 249 (21.8) | 227 (21.3) | 22 (28.9) |  |
| ≥80 | 48 (4.2) | 46 (4.3) | 2 (2.6) |  |
| **Sex** |  |  |  | **0.238** |
| Female | 633 (55.3) | 586 (54.9) | 47 (61.8) |  |
| Male | 511 (44.7) | 482 (45.1) | 29 (38.2) |  |
| **Residence** |  |  |  | **0.560** |
| Urban | 856 (74.8) | 797 (74.6) | 59 (77.6) |  |
| Rural | 288 (25.2) | 271 (25.4) | 17 (22.4) |  |
| **Cancer type** |  |  |  | **0.533** |
| Head and Neck (C00-14, C30-32) | 46 (4.0) | 45 (4.2) | 1 (1.3) |  |
| Upper GI (C15, C16) | 140 (12.2) | 135 (12.6) | 5 (6.6) |  |
| Lower GI (C18-25) | 238 (20.8) | 223 (20.9) | 15 (19.7) |  |
| Lung (C34) | 184 (16.1) | 170 (15.9) | 14 (18.4) |  |
| Skin, mesothelial, and Soft Tissue (C43-44, 46, 49) | 73 (6.4) | 65 (6.1) | 8 (10.5) |  |
| Breast (C50) | 151 (13.2) | 139 (13.0) | 12 (15.8) |  |
| Female Genital Organs (C51-58) | 122 (10.7) | 116 (10.9) | 6 (7.9) |  |
| Prostate (C61) | 49 (4.3) | 45 (4.2) | 4 (5.3) |  |
| Urinary System (C64-68) | 76 (6.6) | 71 (6.6) | 5 (6.6) |  |
| Other | 65 (5.7) | 59 (5.5) | 6 (7.9) |  |
| **Stage at diagnosis** |  |  |  | **0.001** |
| I | 208 (18.2) | 187 (17.5) | 21 (27.6) |  |
| II | 241 (21.1) | 216 (20.2) | 25 (32.9) |  |
| III | 330 (28.8) | 312 (29.2) | 18 (23.7) |  |
| IV | 350 (30.6) | 340 (31.8) | 10 (13.2) |  |
| Unknown | 15 (1.3) | 13 (1.2) | 2 (2.6) |  |
| **Active anticancer treatment** |  |  |  | **0.035** |
| No | 514 (44.9) | 471 (44.1) | 43 (56.6) |  |
| Yes | 630 (55.1) | 597 (55.9) | 33 (43.4) |  |
| **Emphysema** |  |  |  | **0.792** |
| Absent | 1060 (92.7) | 989 (92.6) | 71 (93.4) |  |
| Present | 84 (7.3) | 79 (7.4) | 5 (6.6) |  |
| **AI-detected COVID-19 pneumonia** |  |  |  | **0.68** |
| No | 591 (51.7) | 550 (51.5) | 41 (53.9) |  |
| Yes | 553 (48.3) | 518 (48.5) | 35 (46.1) |  |
| **Coronary artery calcium score category (Agatston units)\*** |  |  |  | **0.126** |
| 0 | 568 (49.7) | 539 (50.5) | 29 (38.2) |  |
| 1-99 | 263 (23.0) | 238 (22.3) | 25 (32.9) |  |
| 100-299 | 139 (12.1) | 129 (12.1) | 10 (13.2) |  |
| ≥300 | 174 (15.2) | 162 (15.2) | 12 (15.8) |  |
| **Epicardial fat volume** |  |  |  | **0.006** |
| <125 mL | 519 (58.6) | 493 (59.8) | 26 (41.9) |  |
| ≥125 mL | 367 (41.4) | 331 (40.2) | 36 (58.1) |  |
| **Bone density** |  |  |  | **0.235** |
| Normal | 402 (35.1) | 382 (35.8) | 20 (26.3) |  |
| Osteopenia | 426 (37.2) | 395 (37.0) | 31 (40.8) |  |
| Osteoporosis | 316 (27.6) | 291 (27.2) | 25 (32.9) |  |

Notes. Data are presented as n (%). P-values were derived from Pearson's chi-squared test. GI = gastrointestinal tract. Coronary artery calcium was quantified using the automated Agatston scoring method and categorized as 0, 1–99, 100–299, and ≥300 Agatston units.

As shown in Table 1, vaccinated patients were significantly older (p=0.019) and had less advanced disease compared to unvaccinated patients. Fewer than half of the vaccinated patients were undergoing active anticancer treatment at the time of CT, compared with 55.9% of unvaccinated patients (p=0.035). No statistically significant differences were found in the distribution of cancer types between the vaccinated and unvaccinated groups. Biomarkers of chronic non-communicable diseases, such as emphysema, coronary artery calcification, and osteopenia/osteoporosis, were generally distributed equally between the two groups. However, the proportion of patients with a high volume of epicardial adipose tissue was significantly greater in the vaccinated group: 36 (58.1%) vs. 331 (40.2%) in the unvaccinated group (p=0.006).

### Overall survival (OS)

During follow-up of the entire cohort, 667 deaths were recorded. Most deaths (567, 85.0%) were attributed to cancer progression, while 41 (6.1%) were attributed to COVID-19 or its complications, and 40 (6.0%) were due to cardiovascular diseases and 19 (2.9%) were due to other or unknown causes.

In the primary landmark cohort (patients alive as of March 1, 2021; N = 1,136), the 1-year and 3-year OS rates were 70.3% (95% CI 67.5–72.8%) and 48.0% (95% CI 45.0–50.9%), respectively. In the sensitivity landmark cohort (patients alive as of November 1, 2021; N = 894), the corresponding OS rates were 89.3% (95% CI 87.0–91.1%) and 61.0% (95% CI 57.7–64.1%).

A significant survival benefit was observed in vaccinated patients. In the primary landmark cohort (March 1, 2021), the 1-year and 3-year overall survival (OS) rates were 98.7% (95% CI 91.0– 99.8%) and 80.3% (95% CI 69.4–87.6%) for vaccinated patients, compared with 68.2% (95% CI 65.3–70.9%) and 45.7% (95% CI 42.6–48.6%) for unvaccinated patients (log-rank p < 0.001; Figure 3A). In the sensitivity landmark cohort (November 1, 2021), the corresponding 1-year and 3-year OS rates were 98.7% (95% CI 91.0–99.8%) and 80.3% (95% CI 69.4–87.6%) for vaccinated patients, compared with 88.4% (95% CI 86.0–90.4%) and 59.2% (95% CI 55.7–62.5%) for unvaccinated patients (log-rank p = 0.001; Figure 3B).

**Figure 3.**
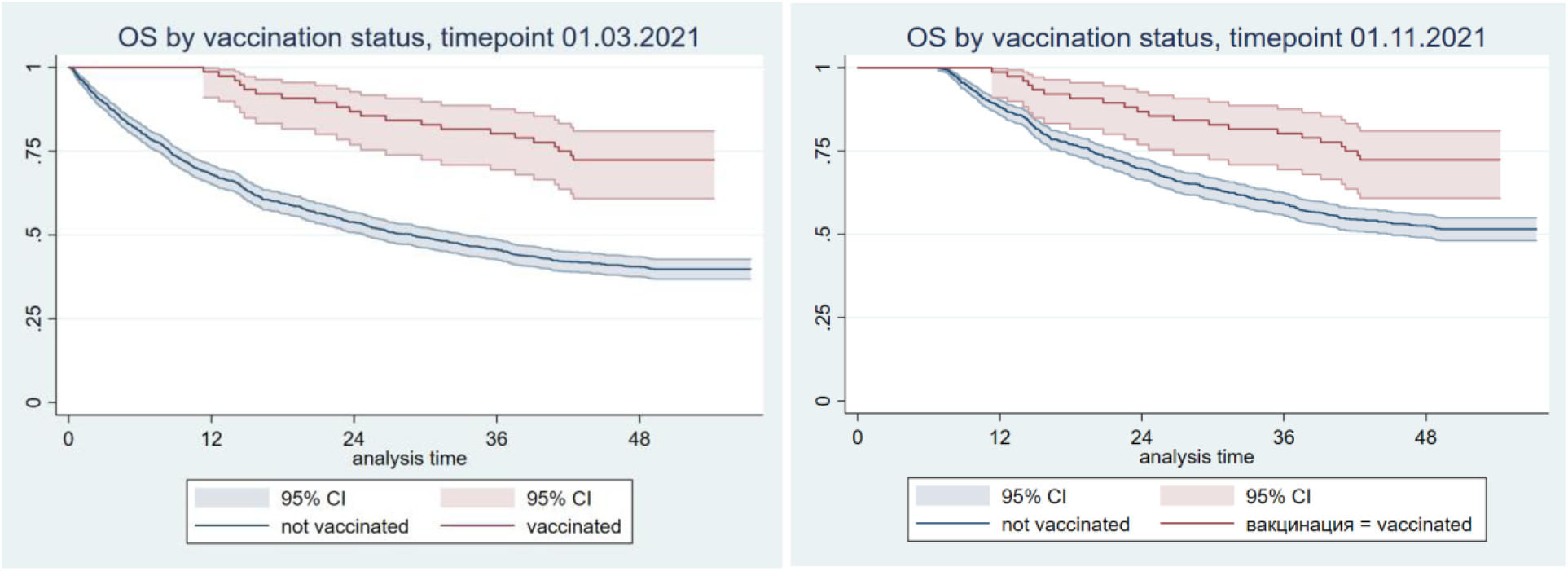
Kaplan-Meier analysis of overall survival by vaccination status. A) Landmark date March 1, 2021, B) Landmark date November 1, 2021. The shaded areas around each curve represent the 95% confidence interval. Log-rank test: (A) p < 0.001; (B) p = 0.001.

Vaccinated patients had a consistently higher probability of survival over time than unvaccinated patients. The results of univariable and multivariable Cox regression analyses of OS according to vaccination status, baseline demographic and cancer characteristics, and AI findings on CT are presented in Table 2.

**Table 2.**
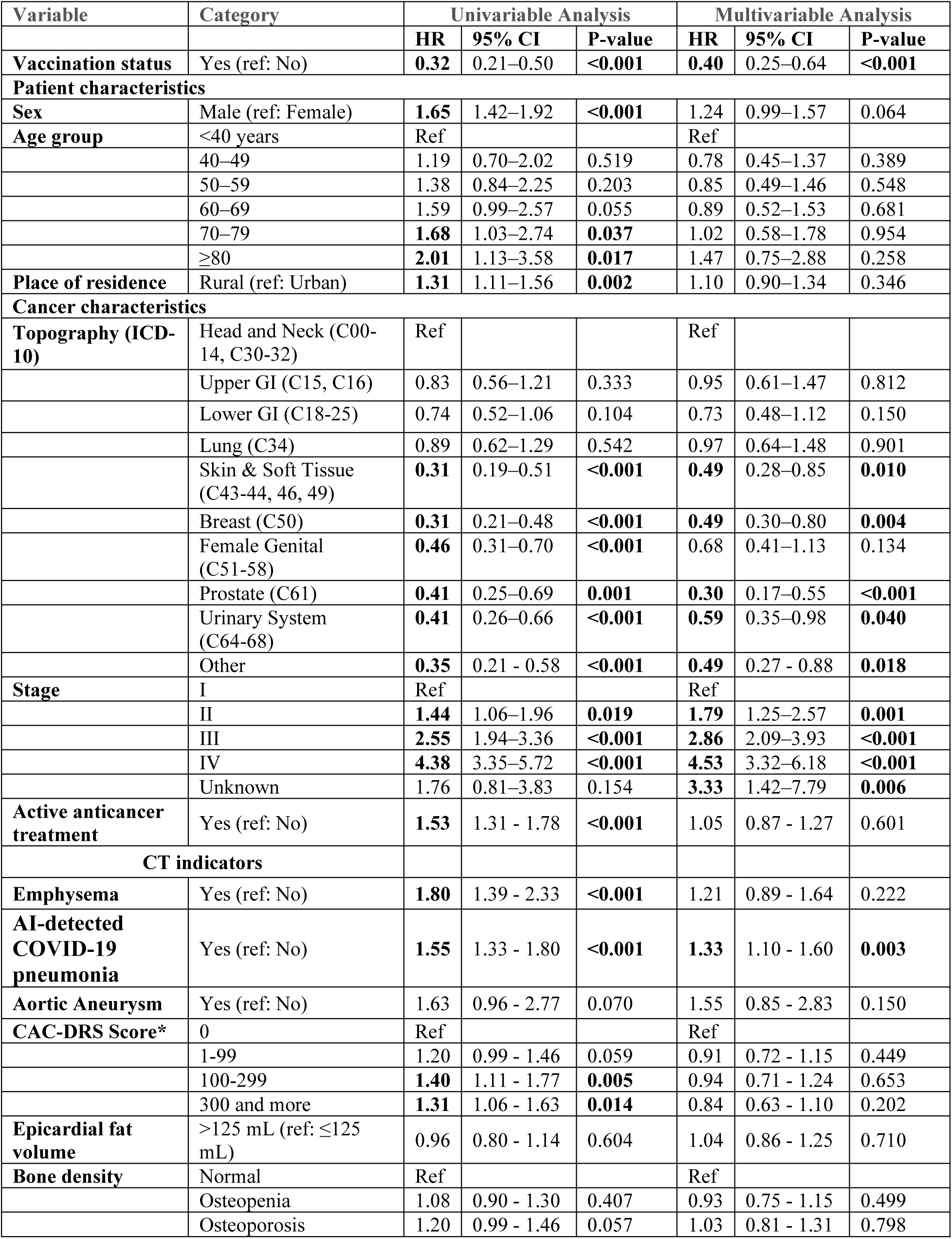
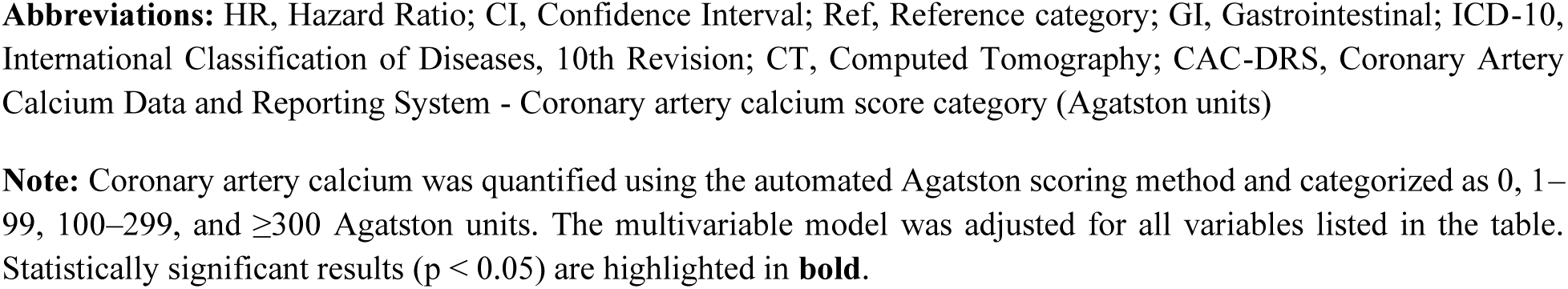
Results of univariable and multivariable Cox regression analysis of overall survival among cancer patients alive as of March 1, 2021, undergoing chest CT in Arkhangelsk, Northwestern Russia, 2020–2021 (N=1,136)

| Variable | Category | Univariable Analysis |  |  | Multivariable Analysis |  |  |
| --- | --- | --- | --- | --- | --- | --- | --- |
|  |  | HR | 95% CI | P-value | HR | 95% CI | P-value |
| **Vaccination status** | Yes (ref: No) | **0.32** | 0.21–0.50 | **<0.001** | **0.40** | 0.25–0.64 | **<0.001** |
| **Patient characteristics** |  |  |  |  |  |  |  |
| **Sex** | Male (ref: Female) | **1.65** | 1.42–1.92 | **<0.001** | 1.24 | 0.99–1.57 | 0.064 |
| **Age group** | <40 years | Ref |  |  | Ref |  |  |
|  | 40–49 | 1.19 | 0.70–2.02 | 0.519 | 0.78 | 0.45–1.37 | 0.389 |
|  | 50–59 | 1.38 | 0.84–2.25 | 0.203 | 0.85 | 0.49–1.46 | 0.548 |
|  | 60–69 | 1.59 | 0.99–2.57 | 0.055 | 0.89 | 0.52–1.53 | 0.681 |
|  | 70–79 | **1.68** | 1.03–2.74 | **0.037** | 1.02 | 0.58–1.78 | 0.954 |
|  | ≥80 | **2.01** | 1.13–3.58 | **0.017** | 1.47 | 0.75–2.88 | 0.258 |
| **Place of residence** | Rural (ref: Urban) | **1.31** | 1.11–1.56 | **0.002** | 1.10 | 0.90–1.34 | 0.346 |
| **Cancer characteristics** |  |  |  |  |  |  |  |
| **Topography (ICD-10)** | Head and Neck (C00–14, C30–32) | Ref |  |  | Ref |  |  |
|  | Upper GI (C15, C16) | 0.83 | 0.56–1.21 | 0.333 | 0.95 | 0.61–1.47 | 0.812 |
|  | Lower GI (C18–25) | 0.74 | 0.52–1.06 | 0.104 | 0.73 | 0.48–1.12 | 0.150 |
|  | Lung (C34) | 0.89 | 0.62–1.29 | 0.542 | 0.97 | 0.64–1.48 | 0.901 |
|  | Skin & Soft Tissue (C43–44, 46, 49) | **0.31** | 0.19–0.51 | **<0.001** | **0.49** | 0.28–0.85 | **0.010** |
|  | Breast (C50) | **0.31** | 0.21–0.48 | **<0.001** | **0.49** | 0.30–0.80 | **0.004** |
|  | Female Genital (C51–58) | **0.46** | 0.31–0.70 | **<0.001** | 0.68 | 0.41–1.13 | 0.134 |
|  | Prostate (C61) | **0.41** | 0.25–0.69 | **0.001** | **0.30** | 0.17–0.55 | **<0.001** |
|  | Urinary System (C64–68) | **0.41** | 0.26–0.66 | **<0.001** | **0.59** | 0.35–0.98 | **0.040** |
|  | Other | **0.35** | 0.21–0.58 | **<0.001** | **0.49** | 0.27–0.88 | **0.018** |
| **Stage** | I | Ref |  |  | Ref |  |  |
|  | II | **1.44** | 1.06–1.96 | **0.019** | **1.79** | 1.25–2.57 | **0.001** |
|  | III | **2.55** | 1.94–3.36 | **<0.001** | **2.86** | 2.09–3.93 | **<0.001** |
|  | IV | **4.38** | 3.35–5.72 | **<0.001** | **4.53** | 3.32–6.18 | **<0.001** |
|  | Unknown | 1.76 | 0.81–3.83 | 0.154 | **3.33** | 1.42–7.79 | **0.006** |
| **Active anticancer treatment** | Yes (ref: No) | **1.53** | 1.31–1.78 | **<0.001** | 1.05 | 0.87–1.27 | 0.601 |
| **CT indicators** |  |  |  |  |  |  |  |
| **Emphysema** | Yes (ref: No) | **1.80** | 1.39–2.33 | **<0.001** | 1.21 | 0.89–1.64 | 0.222 |
| **AI-detected COVID-19 pneumonia** | Yes (ref: No) | **1.55** | 1.33–1.80 | **<0.001** | **1.33** | 1.10–1.60 | **0.003** |
| **Aortic Aneurysm** | Yes (ref: No) | 1.63 | 0.96–2.77 | 0.070 | 1.55 | 0.85–2.83 | 0.150 |
| **CAC-DRS Score\*** | 0 | Ref |  |  | Ref |  |  |
|  | 1–99 | 1.20 | 0.99–1.46 | 0.059 | 0.91 | 0.72–1.15 | 0.449 |
|  | 100–299 | **1.40** | 1.11–1.77 | **0.005** | 0.94 | 0.71–1.24 | 0.653 |
|  | 300 and more | **1.31** | 1.06–1.63 | **0.014** | 0.84 | 0.63–1.10 | 0.202 |
| **Epicardial fat volume** | >125 mL (ref: ≤125 mL) | 0.96 | 0.80–1.14 | 0.604 | 1.04 | 0.86–1.25 | 0.710 |
| **Bone density** | Normal | Ref |  |  | Ref |  |  |
|  | Osteopenia | 1.08 | 0.90–1.30 | 0.407 | 0.93 | 0.75–1.15 | 0.499 |
|  | Osteoporosis | 1.20 | 0.99–1.46 | 0.057 | 1.03 | 0.81–1.31 | 0.798 |

**Abbreviations:** HR, Hazard Ratio; CI, Confidence Interval; Ref, Reference category; GI, Gastrointestinal; ICD-10, International Classification of Diseases, 10th Revision; CT, Computed Tomography; CAC-DRS, Coronary Artery Calcium Data and Reporting System - Coronary artery calcium score category (Agatston units)
**Note:** Coronary artery calcium was quantified using the automated Agatston scoring method and categorized as 0, 1–99, 100–299, and $\geq 300$ Agatston units. The multivariable model was adjusted for all variables listed in the table. Statistically significant results ( $p < 0.05$ ) are highlighted in **bold**.

Univariable analysis revealed that vaccination was associated with a 68% lower hazard of all-cause mortality (HR 0.32, 95% CI 0.21–0.50). Adjustment for available confounders in the multivariable model only slightly attenuated this association, which remained significant ratio (HR 0.40; 95% CI, 0.25–0.64).

Other independent predictors of poor prognosis in the multivariable model included advanced disease stage (adjusted HRs ranging from 1.79 for stage II to 4.53 for stage IV, compared with stage I) and AI-detected COVID-19 pneumonia on chest CT (HR 1.33, 95% CI 1.10–1.60). Conversely, patients with cancers of the skin and soft tissues, breast, prostate, and urinary system demonstrated a more favorable OS.

Several factors significantly associated with poorer survival in univariable analysis—male sex, older age, rural residence, and active anticancer treatment—lost statistical significance after multivariable adjustment.

Furthermore, the adverse prognostic impact of AI-quantified emphysema (unadjusted HR 1.80, 95% CI 1.39–2.33) and a high coronary calcium burden (CAC-DRS score >100, unadjusted HR 1.31–1.40) on chest CT was no longer significant after multivariable adjustment. Low bone density on CT was not associated with worse OS.

In a sensitivity analysis restricted to patients alive as of November 1, 2021the unadjusted HR for vaccination was 0.47 (95% CI 0.30–0.73). After adjustment for all covariates listed in Table 2, the HR was 0.56 (95% CI 0.34–0.92).

## Discussion

This population-based cohort study, leveraging integrated data from a regional cancer registry, AI-powered CT analysis, and official vaccination records, provides critical insights into COVID-19 vaccination coverage and its association with survival among cancer patients in Northwestern Russia. Our two principal findings are: first, an alarmingly low vaccination rate of 6.6% in this high-risk cohort during the study period; and second, a robust and independent association between vaccination and a significant reduction in all-cause mortality, with vaccinated patients in the primary analysis (March 1, 2021) experiencing a 60% lower hazard of death (adjusted HR 0.40, 95% CI 0.25–0.64). In a sensitivity analysis restricted to patients alive as of November 1, 2021, the adjusted HR was 0.56 (95% CI 0.34–0.92), showing a consistent, but attenuated survival benefit, likely due to shorter follow-up in this later cohort.

The attenuation of the survival benefit observed in the later cohort (November 1, 2021) compared with the primary analysis (March 1, 2021) is likely multifactorial. While the smaller sample size and shorter follow-up in this sensitivity analysis reduce statistical power to detect the full effect, another important factor is the changing landscape of the pandemic itself. By late 2021, the Delta variant remained dominant, and the Omicron variant, which was associated with less severe disease in vaccinated and previously infected individuals, emerged shortly thereafter. Consequently, the baseline mortality risk from COVID-19 among unvaccinated cancer patients may have been lower in the later period, narrowing the survival gap between vaccinated and unvaccinated groups. This hypothesis is consistent with the OnCovid study findings, which reported less severe outcomes during the Omicron wave compared with earlier phases, even among unvaccinated patients with cancer [10]. The persistence of a statistically significant survival benefit across both landmark dates, despite this potential reduction in intrinsic viral virulence, underscores the robustness of the association between vaccination and improved survival in this vulnerable population.

The strikingly low vaccination coverage observed in our cohort (6.6%) stands in stark contrast to reported figures from other regions. During the initial phase of the vaccination rollout, Russia reported one of the lowest vaccination rates among the general population [18]. Among cancer patients, this rate was markedly below both the 59.0% pooled acceptance rate described in a global meta-analysis [11] and the 33.3% coverage observed in European cancer patients undergoing active treatment during the Omicron phase [10]. These findings highlight considerable disparities and pronounced heterogeneity (I² = 99%) in vaccination uptake among cancer patients globally, as outlined by Prabani et al. [11], raising important public health considerations within the Russian healthcare system. The reasons for this low uptake are likely multifactorial, encompassing vaccine hesitancy, concerns among clinicians and patients regarding interactions between vaccines and cancer therapy, and potential logistical barriers within the healthcare system.

Our data suggest a possible selection bias wherein vaccinated patients were significantly more likely to have earlier-stage disease and were less frequently on active treatment, indicating that clinicians and patients may have been more inclined towards vaccination during periods of stable disease [19]. This hypothesis aligns with the observed demographic profile of the vaccinated group, who were older -a factor associated with higher vaccine acceptance in the general population in many countries [20; 21] due to perceived higher risk. Notably, vaccinated patients also had a significantly higher proportion of elevated epicardial adipose tissue volume (p=0.006), a biomarker associated with cardiovascular risk, which may reflect a greater awareness of comorbidities and consequently higher vaccination acceptance.

Despite this low coverage, the survival benefit associated with vaccination was substantial. The magnitude of the protective effect-a three-fold reduction in mortality risk in univariate analysis (HR 0.32) that remained a 2.5-fold reduction after multivariate adjustment (HR 0.40)-is noteworthy. This finding is consistent with the established international consensus that COVID-19 vaccination is associated with improved survival in cancer patients [3; 4; 9]. Our results extend this evidence base to the Russian population and to the specific context of adenovirus-vectored vaccines (Sputnik V and Sputnik Light), supporting their real-world effectiveness in a clinical oncology setting. The survival curves show an early and sustained separation, suggesting that vaccination was particularly important in preventing deaths from COVID-19 during the prevalent phases of the pandemic covered by our CT data acquisition.

The multivariate analysis further validated other key predictors of mortality. As expected, advanced cancer stage was the strongest predictor of poor outcome, with adjusted HRs ranging from 1.79 (stage II) to 4.53 (stage IV) compared with stage I. Importantly, the presence of radiological signs of COVID-19 pneumonia on CT, as identified by the AI algorithm, remained an independent predictor of increased mortality (adjusted HR 1.32, 95% CI 1.09–1.59), corroborating previous findings that asymptomatic or pre-symptomatic COVID-19 in cancer patients carries a significant risk of adverse outcomes [12] and reinforcing the value of AI-assisted CT analysis in risk stratification. The consistency of this finding across both univariate and multivariate models highlights the prognostic importance of even incidental radiological findings suggestive of COVID-19.

We used a certified medical device (RZN 2024/22895) capable of automated multi-organ phenotyping from chest CT scans. Its architecture enabled quantitative assessment of COVID-19-associated pulmonary involvement while simultaneously extracting cardiovascular and metabolic biomarkers including coronary artery calcification, epicardial adipose tissue volume, aortic diameters, and vertebral bone mineral density, thus generating a comprehensive dataset for survival modeling in oncology. By ensuring reproducibility and standardization across large imaging cohorts, the IRA Labs system transformed routine diagnostic scans into a multiparametric prognostic resource, representing a significant advance in both clinical and research applications for cancer populations under pandemic conditions. This approach underscores the potential of certified AI solutions to facilitate robust, scalable, and reproducible risk stratification workflows in oncological practice [15; 16].

The use of reproducible AI-processed data offers several advantages. Fixed model versions and unified threshold protocols produce consistent results when applied to identical datasets, which helps to reduce methodological error and interobserver variability, and consequently increases statistical power. Secondly, standardized quantitative metrics allow for valid subgroup comparisons, which is particularly crucial when analyzing cancer patients with COVID-19, and facilitate external replication of findings. Finally, multi-organ phenotyping from a single CT scan, including lungs, vessels, bones, and adipose tissue, enables comprehensive adjustment for confounding factors and supports the development of integrated risk models, enhancing the clinical applicability of our conclusions.

Several factors significant in univariable analysis, such as active cancer treatment, male sex, and the presence of emphysema or high coronary calcium, lost their independent prognostic value in the multivariable model, suggesting their effect on overall mortality is mediated through or confounded by other variables, primarily cancer stage and vaccination status. The loss of significance for active treatment after adjustment is noteworthy. This attenuation may reflect confounding by stage and other factors, but it is also consistent with the hypothesis that vaccination mitigates some of the excess risk associated with active anticancer therapy. This is consistent with a retrospective U.S. study showing that while vaccine effectiveness was 85% in patients without recent systemic therapy, it dropped to 57% in those receiving chemotherapy. Nonetheless, about 10% of vaccinated patients with breakthrough infections still died—a rate similar to that of unvaccinated infected patients [22]. These findings support the recommendation for vaccination regardless of treatment status.

The quality of data from the PBCR of the Arkhangelsk Region and the Nenets Autonomous Okrug meets international standards, which has been repeatedly confirmed in international studies, including Cancer Incidence in Five Continents, Volume XII [23]. According to the data from this study, the percentage of morphologically verified cases (MV%) for all cancers, excluding non-melanoma skin cancer (C00-C96, excl. C44), registered in the coverage area of the PBCR of the Arkhangelsk and Nenets Autonomous Okrug, reaches 84.2% in men and 86.3% in women. The proportion of cases recorded only based on death certificates (DCO%) among men is 8.5%, and among women it is 7.3%. This high level of morphological verification indicates the reliability of these cancer registries. Additionally, the relatively low DCO% demonstrates the high completeness of cancer case recording in the registries.

### Limitations

This study has several limitations. First, its observational design cannot definitively establish causality, although the strength and consistency of the association with adjustment for key confounders prove the inference. Second, although overall survival using all-cause mortality serves as a rigorous endpoint, it may obscure the specific impact of vaccination on COVID-19-related mortality. However, given that COVID-19 can exacerbate other conditions leading to death, OS is a comprehensive measure of the overall protective benefit of vaccination during the pandemic. Third, the generalizability of the vaccination coverage rate may be limited, as our cohort consisted of patients undergoing CT scanning, who may differ from the general oncology population. Finally, the SNILS-based data linkage, while accurate, resulted in the exclusion of a substantial number of patients due to missing identifiers, potentially introducing selection bias.

## Conclusion and Implications

In conclusion, this study reveals two contrasting messages. First, COVID-19 vaccination coverage among cancer patients in a Russian region was critically low signaling a need for targeted interventions to address vaccine hesitancy and improve access in this vulnerable group - an issue of particular importance for preparedness for future pandemics. Second, despite the low coverage, vaccination was associated with a substantial survival benefit with a 60% lower hazard of all-cause mortality (adjusted HR 0.40; 95% CI, 0.25–0.64). The integration of AI-powered CT analysis further highlights the potential utility of advanced imaging biomarkers for comprehensive risk assessment in oncology patients.

These findings suggest that public health authorities and oncologists should prioritize COVID-19 vaccination as part of standard cancer care during future pandemics. Future efforts should focus on understanding and overcoming the barriers to vaccination in this population to ensure that the potential survival benefit observed in the study setting is realized for all cancer patients. Further research in larger, multicenter cohorts is warranted to corroborate these findings and to explore strategies for improving vaccine uptake.

## Data Availability

All data produced in the present study are available upon reasonable request to the authors

## Author’s Contribution

I. Conception and design: AAD, VAG, MYV
II. Administrative support: DVB, VAG, MYV
III. Provision of study materials or patients: AAD, MAB, DVB, VYC, VAG, MYV
IV. Collection and assembly of data: AAD, MAB, VYC, VAG, MYV
V. Data analysis and interpretation: AAD, MAB, DVB, AAM, VYC, VAG, AMG, MYV
VI. Manuscript writing: All authors
VII. Final approval of manuscript: All authors

## Acknowledgments

The authors express their gratitude to the staff of the Radiology Department at the Arkhangelsk Clinical Oncology Dispensary for their intensive work on data collection during the COVID-19 pandemic. The authors also thank the staff of the Population Cancer Registry of the Arkhangelsk Region and the Nenets Autonomous Okrug for collecting, analyzing, and interpreting data on patients with malignant neoplasms from two regions of the Russian Federation for more than two decades, which is truly unique in the country. In addition, the authors thank the staff of the Medical Information and Analytical Center for their assistance in enriching the study database with vaccination information.

## Footnote

a. Conflicts of Interest: Valeria Yu. Chernina is a Head of the Clinical Evaluation Department of the LLC IRA Labs, Viktor A. Gombolevsky is an Advisor of the LLC IRA Labs. Other authors declare no conflicts of interest.
b. Financial Disclose: The study was conducted using funds and resources from IRA Labs.
c. Ethical statement: The study was approved by the Local Ethics Committee of the Northern State Medical University (Protocol No. 07/10-238, 2023). The study conforms to the provisions of in accordance with the Helsinki Declaration as revised in 2013, available at: http://www.wma.net/en/30publications/10policies/b3/%20index.html.

Authors are accountable for all aspects of the work (including full data access, integrity of the data and the accuracy of the data analysis) in ensuring that questions related to the accuracy or integrity of any part of the work are appropriately investigated and resolved.

## Supplementary material

### Appendix 1

Characteristics of all evaluated AI-derived imaging biomarkers and their performance in independent external validation within the Innovative Technologies Experiment (mosmed.ai), reported as ROC AUC, sensitivity, specificity, and accuracy.

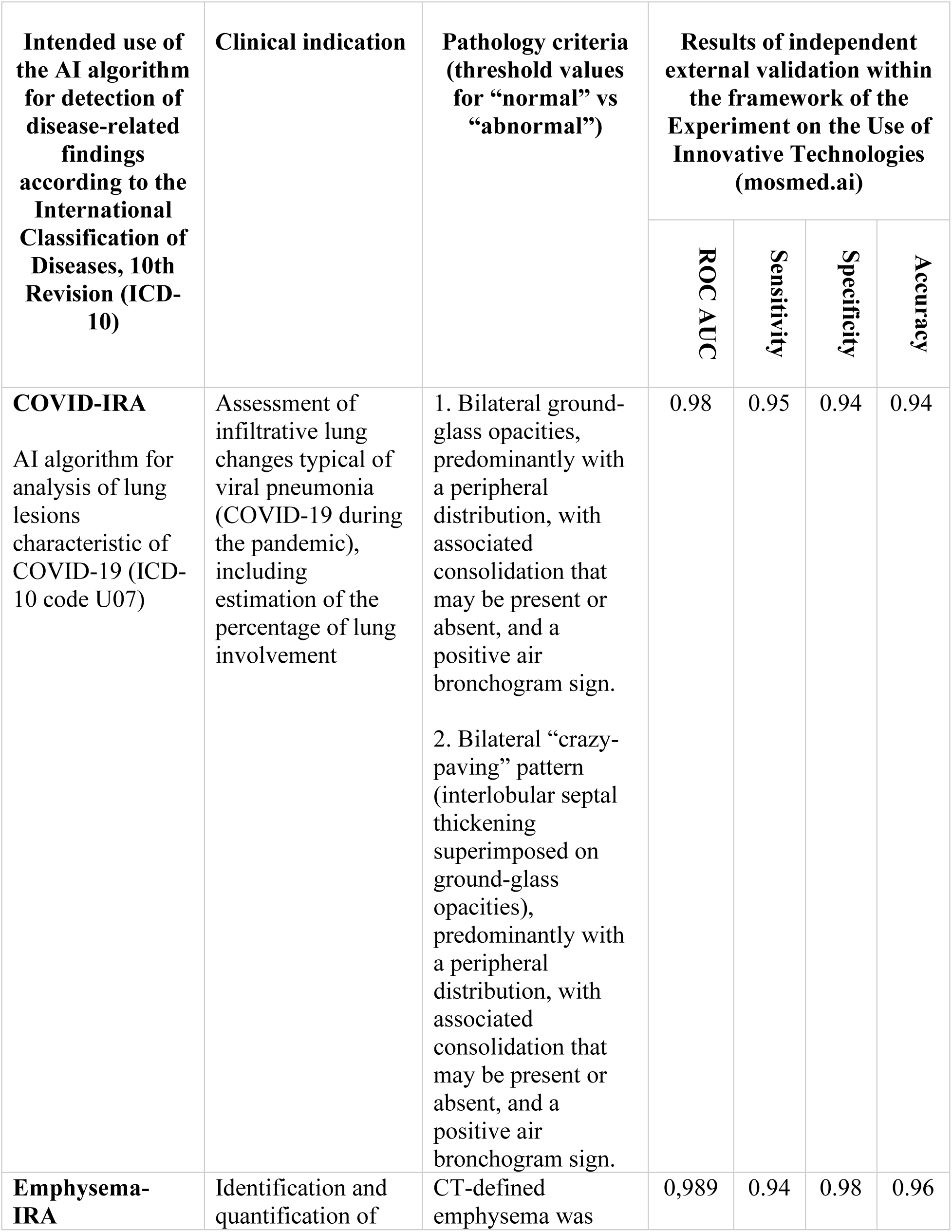
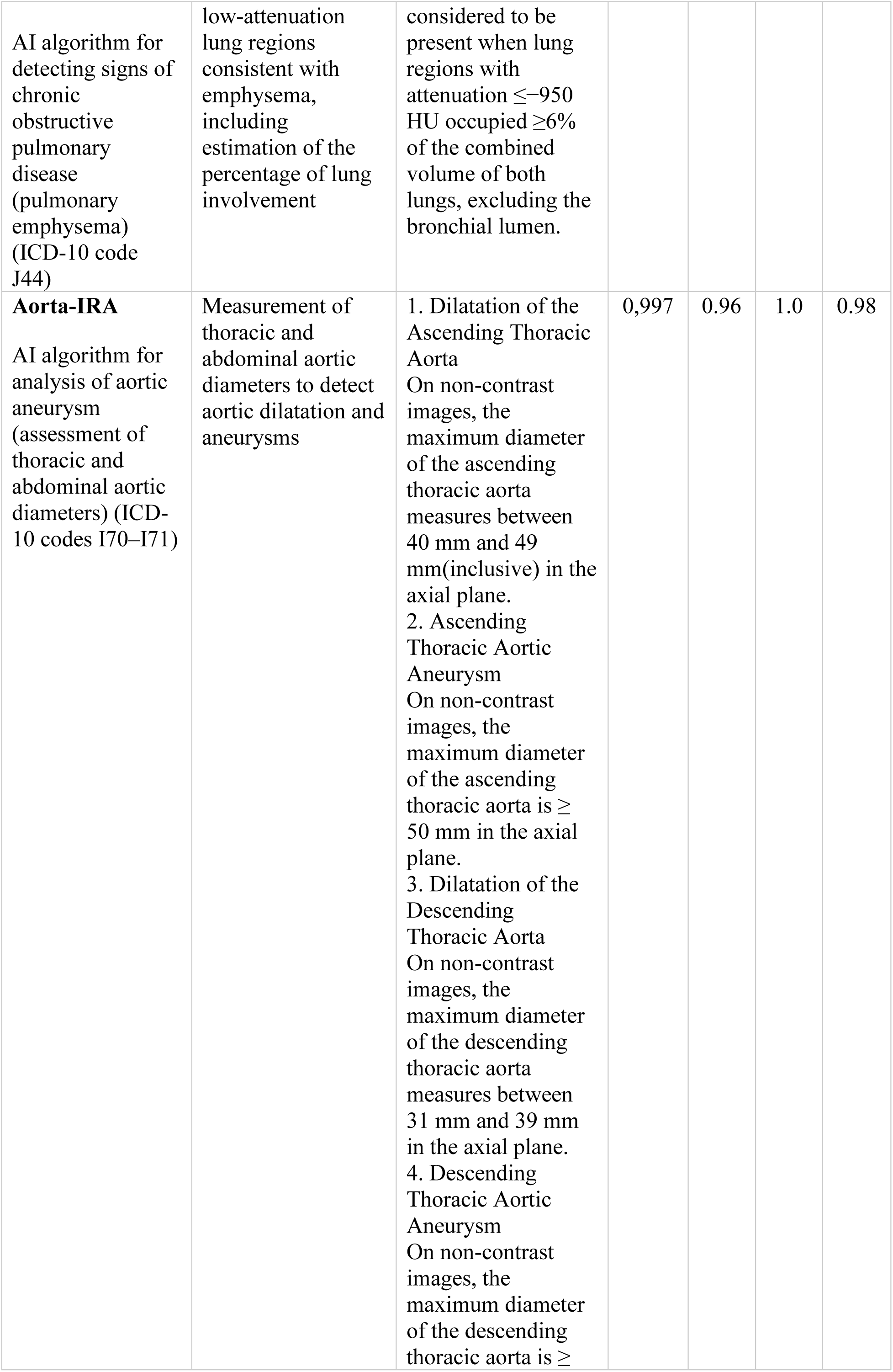
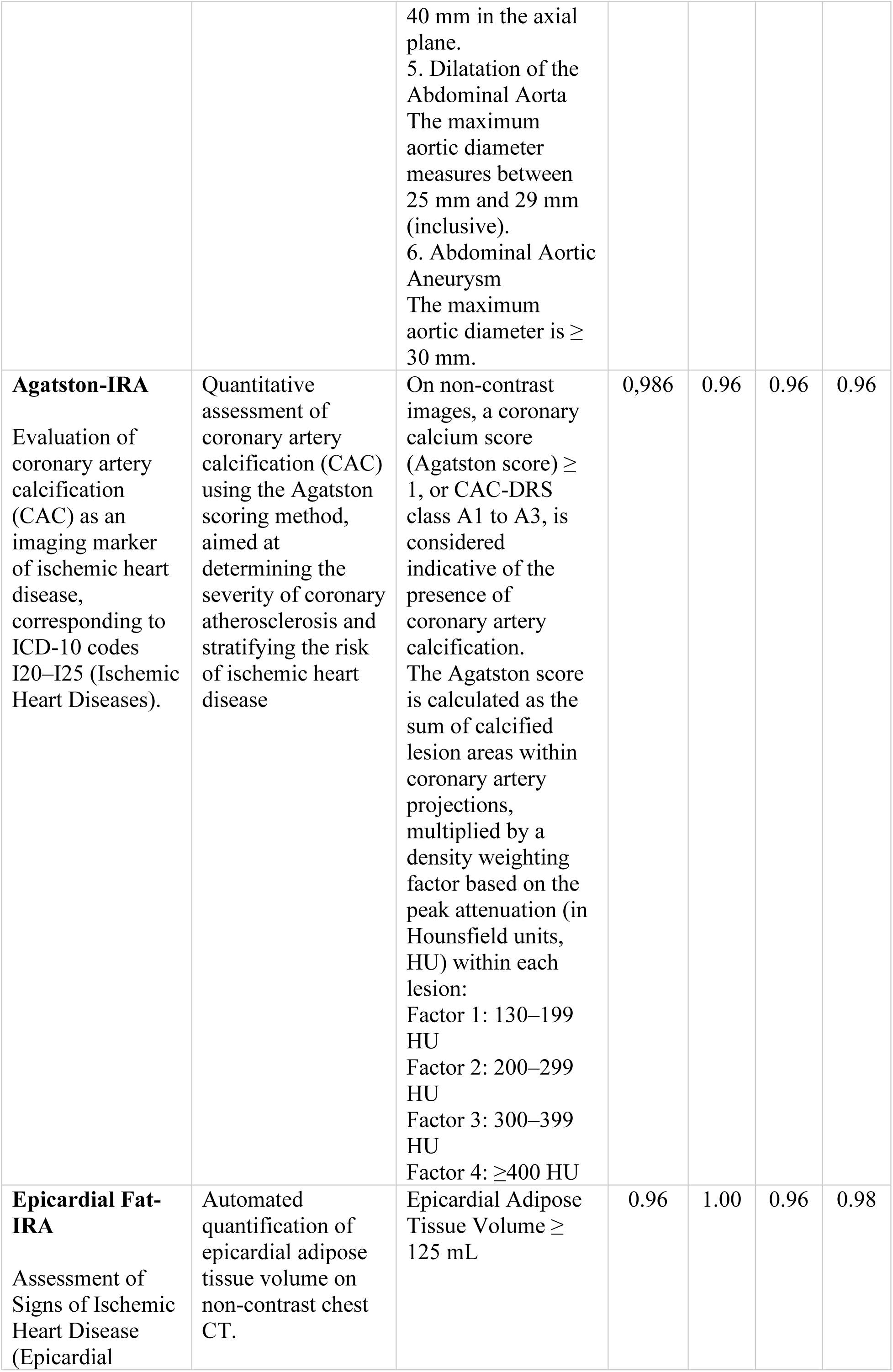
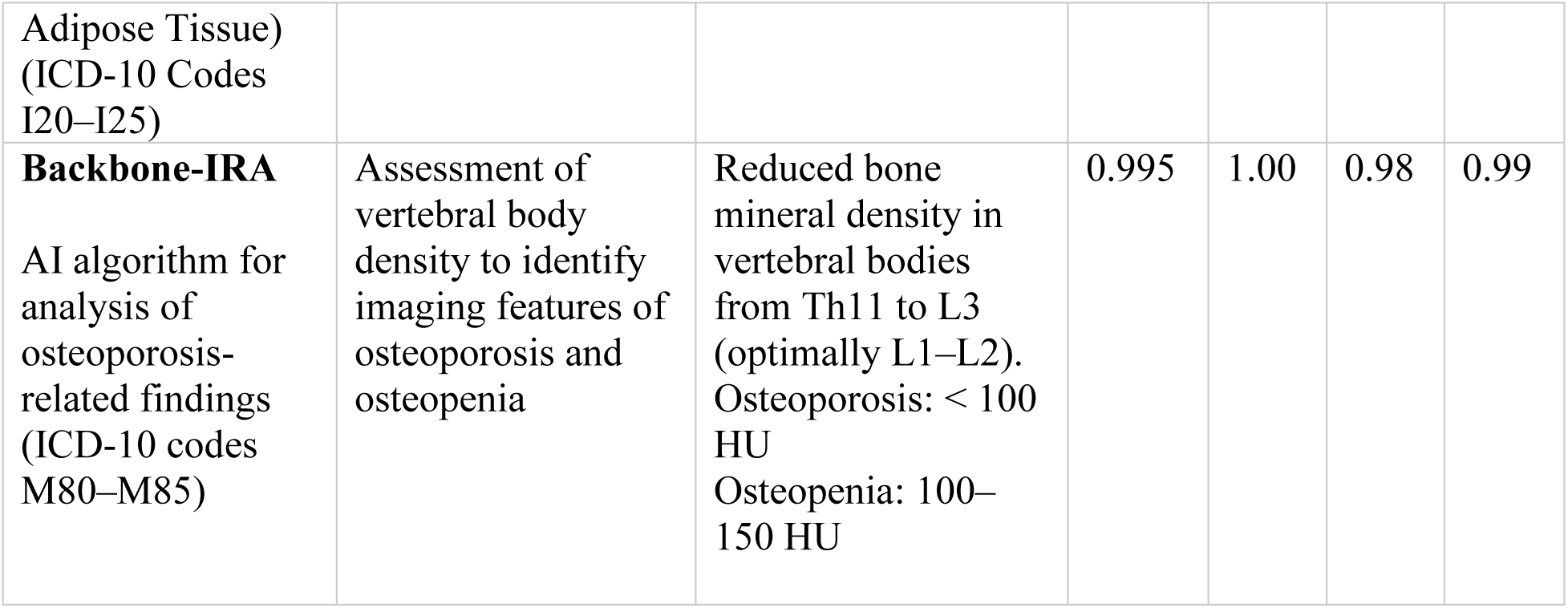

## Notes

### Author Declarations

The study was approved by the Local Ethics Committee of the Northern State Medical University (Protocol No. 07/10-238, 2023)

### Funding

- Intelligent Radiology Assistance Laboratories LLC (IRA LABS)

## References

1. World Health Organization. WHO COVID-19 deaths dashboard [Internet]. Geneva: WHO; [cited 2026 Aug 20]. Available from: https://data.who.int/dashboards/covid19/deaths?n=o

2. World Health Organization. Global excess deaths associated with COVID-19 (modelled estimates) (Russian Federation) [Internet]. Geneva: WHO; [cited 2026 Aug 20]. Available from: https://www.who.int/data/sets/global-excess-deaths-associated-with-covid-19-modelled-estimates

3. Kuderer NM, Choueiri TK, Shah DP, et al. Clinical impact of COVID-19 on patients with cancer (CCC19): a cohort study. Lancet. 2020;395:1907–1918. doi:10.1016/S0140-6736(20)31187-9.

4. Grivas P, Khaki AR, Wise-Draper TM, et al. Association of clinical factors and recent anticancer therapy with COVID-19 severity among patients with cancer: a report from the COVID-19 and Cancer Consortium. Ann Oncol. 2021;32:787–800. doi:10.1016/j.annonc.2021.02.024.

5. World Health Organization. WHO SAGE roadmap for prioritizing uses of COVID-19 vaccines in the context of limited supply [Internet]. Geneva: WHO; 2020 Oct 14 [cited 2026 Aug 24]. Available from: https://www.who.int/publications/i/item/who-sage-roadmap-for-prioritizing-uses-of-covid-19-vaccines-in-the-context-of-limited-supply

6. European Centre for Disease Prevention and Control. Overview of EU/EEA country recommendations on COVID-19 vaccination with a focus on the elderly and medically vulnerable [Internet]. Stockholm: ECDC; 2021 Feb 4 [cited 2026 Aug 24]. Available from: https://www.ecdc.europa.eu/en/publications-data/overview-eueea-country-recommendations-covid-19-vaccination-focus-elderly-and-medically-vulnerable

7. Ministry of Health of the Russian Federation. On the procedure for vaccination against the novel coronavirus infection (COVID-19) of adult citizens [Internet]. Moscow: Ministry of Health of the Russian Federation; 2020 Dec 21 [cited 2026 Aug 24]. Available from: https://static-0.minzdrav.gov.ru/system/attachments/attaches/000/054/596/original/21122020_M3_COVID-19.pdf (in Russian).

8. Song Q, Bates B, Shao YR, et al. Risk and outcome of breakthrough COVID-19 infections in vaccinated patients with cancer: real-world evidence from the National COVID Cohort Collaborative. J Clin Oncol. 2022;40(13):1414–1427. doi:10.1200/JCO.21.02419.

9. Schmidt AL, Labaki C, Hsu CY, et al. COVID-19 vaccination and breakthrough infections in patients with cancer. Ann Oncol. 2022;33(3):340–346. doi:10.1016/j.annonc.2021.12.006.

10. Pinato DJ, Aguilar-Company J, Ferrante D, et al. Outcomes of the SARS-CoV-2 omicron (B.1.1.529) variant outbreak among vaccinated and unvaccinated patients with cancer in Europe: results from the retrospective, multicentre, OnCovid registry study. Lancet Oncol. 2022;23(7):865–875. doi:10.1016/S1470-2045(22)00273-X.

11. Prabani KIP, Weerasekara I, Damayanthi HDWT. COVID-19 vaccine acceptance and hesitancy among patients with cancer: a systematic review and meta-analysis. Public Health. 2022;212:66–75. doi:10.1016/j.puhe.2022.09.001.

12. Dai M, Liu D, Liu M, et al. Patients with cancer appear more vulnerable to SARS-CoV-2: a multicenter study during the COVID-19 outbreak. Cancer Discov. 2020;10(6):783–791. doi:10.1158/2159-8290.CD-20-0422.

13. Dyachenko AA, Bogdanov MA, Bogdanov DV, et al. Prognostic role of COVID-19 pneumonia signs and other CT-biomarkers for survival in patients with malignant neoplasms: the ARILUS project. medRxiv [Preprint]. 2025 Nov 17. doi:10.1101/2025.11.14.25338295.

14. Valkov MY, Grjibovski AM, Kudryavtsev AV, et al. Using artificial intelligence to predict and prevent non-cancer mortality in patients with cancer: ARILUS study protocol. Ekologiya Cheloveka (Human Ecology). 2024;31(4):314–330. doi:10.17816/humeco635357.

15. Morozov SP, Gombolevskiy VA, Chernina VY, et al. Prediction of lethal outcomes in COVID-19 cases based on chest computed tomography results. Tuberculosis and Lung Diseases. 2020;98(6):7–14. doi:10.21292/2075-1230-2020-98-6-7-14. (in Russian).

16. Vasiliev YA, et al., editors. Computer vision in radiology: the first stage of the Moscow experiment. 2nd ed., rev. and exp. Moscow: Publishing Solutions; 2023. 376 p. (in Russian).

17. Cox DR. Regression models and life-tables. J R Stat Soc Series B Stat Methodol. 1972;34(2):187–220.

18. Sallam M. COVID-19 vaccine hesitancy worldwide: a concise systematic review of vaccine acceptance rates. Vaccines (Basel). 2021;9(2):160. doi:10.3390/vaccines9020160.

19. Shahid Z, Patrick AL, Wallander ML, et al. COVID-19 vaccine uptake trends in SARS-CoV-2 previously infected cancer patients. Vaccine X. 2023;14:100289. doi:10.1016/j.jvacx.2023.100289.

20. Griffiths EA, Segal BH, Alwan LM, et al. COVID-19 vaccination in patients with cancer and SARS-CoV-2 infection in 2020: results from the ASCO Registry. JCO Oncol Pract. 2023;19(7):457–459. doi:10.1200/OP.22.00825.

21. Giesen N, Sprute R, Rüthrich M, et al. 2021 update of the AGIHO guideline on evidence-based management of COVID-19 in patients with cancer regarding diagnostics, viral shedding, vaccination and therapy. Eur J Cancer. 2021;147:154–160. doi:10.1016/j.ejca.2021.01.033.

22. Wu JT, La J, Branch-Elliman W, et al. Association of COVID-19 vaccination with SARS-CoV-2 infection in patients with cancer: a US nationwide Veterans Affairs study. JAMA Oncol. 2022;8:281–286. doi:10.1001/jamaoncol.2021.5771.

23. Bray F, Colombet M, Aitken JF, et al., editors. Cancer incidence in five continents. Vol. XII. IARC Scientific Publication No. 169. Lyon: International Agency for Research on Cancer; 2024. Available from: https://publications.iarc.who.int/641/

