## Appendix 1. for "COVID-19 vaccination and survival of cancer patients in Northwestern Russia: a historical registry-based cohort study"

**Supplementary material**

**Appendix 1.**

Characteristics of all evaluated AI-derived imaging biomarkers and their performance in independent external validation within the Innovative Technologies Experiment (mosmed.ai), reported as ROC AUC, sensitivity, specificity, and accuracy.

| **Intended use of the AI algorithm for detection of disease-related findings according to the International Classification of Diseases, 10th Revision (ICD-10)** | **Clinical indication** | **Pathology criteria (threshold values for "normal" vs "abnormal")** | **Results of independent external validation within the framework of the Experiment on the Use of Innovative Technologies (mosmed.ai)** | | | |
|---|---|---|---|---|---|---|
| | | | **ROC AUC** | **Sensitivity** | **Specificity** | **Accuracy** |
| **COVID-IRA**<br><br>AI algorithm for analysis of lung lesions characteristic of COVID-19 (ICD-10 code U07) | Assessment of infiltrative lung changes typical of viral pneumonia (COVID-19 during the pandemic), including estimation of the percentage of lung involvement | 1. Bilateral ground-glass opacities, predominantly with a peripheral distribution, with associated consolidation that may be present or absent, and a positive air bronchogram sign.<br><br>2. Bilateral "crazy-paving" pattern (interlobular septal thickening superimposed on ground-glass opacities), predominantly with a peripheral distribution, with | 0.98 | 0.95 | 0.94 | 0.94 |

| | | | | | | |
|---|---|---|---|---|---|---|
| | | associated consolidation that may be present or absent, and a positive air bronchogram sign. | | | | |
| **Emphysema-IRA**<br><br>AI algorithm for detecting signs of chronic obstructive pulmonary disease (pulmonary emphysema) (ICD-10 code J44) | Identification and quantification of low-attenuation lung regions consistent with emphysema, including estimation of the percentage of lung involvement | CT-defined emphysema was considered to be present when lung regions with attenuation ≤−950 HU occupied ≥6% of the combined volume of both lungs, excluding the bronchial lumen. | 0,989 | 0.94 | 0.98 | 0.96 |
| **Aorta-IRA**<br><br>AI algorithm for analysis of aortic aneurysm (assessment of thoracic and abdominal aortic diameters) (ICD-10 codes I70–I71) | Measurement of thoracic and abdominal aortic diameters to detect aortic dilatation and aneurysms | 1. Dilatation of the Ascending Thoracic Aorta<br><br>On non-contrast images, the maximum diameter of the ascending thoracic aorta measures between 40 mm and 49 mm(inclusive) in the axial plane.<br><br>2. Ascending Thoracic Aortic Aneurysm<br><br>On non-contrast images, the maximum diameter of the ascending thoracic aorta is ≥ | 0,997 | 0.96 | 1.0 | 0.98 |

| | | |
|---|---|---|
| | | 50 mm in the axial plane.<br><br>3. Dilatation of the Descending Thoracic Aorta<br><br>On non-contrast images, the maximum diameter of the descending thoracic aorta measures between 31 mm and 39 mm in the axial plane.<br><br>4. Descending Thoracic Aortic Aneurysm<br><br>On non-contrast images, the maximum diameter of the descending thoracic aorta is ≥ 40 mm in the axial plane.<br><br>5. Dilatation of the Abdominal Aorta<br><br>The maximum aortic diameter measures between 25 mm and 29 mm (inclusive).<br><br>6. Abdominal Aortic Aneurysm<br><br>The maximum aortic diameter is ≥ 30 mm. |

| | | | | | | |
|---|---|---|---|---|---|---|
| **Agatston-IRA**<br><br>Evaluation of coronary artery calcification (CAC) as an imaging marker of ischemic heart disease, corresponding to ICD-10 codes I20–I25 (Ischemic Heart Diseases). | Quantitative assessment of coronary artery calcification (CAC) using the Agatston scoring method, aimed at determining the severity of coronary atherosclerosis and stratifying the risk of ischemic heart disease | On non-contrast images, a coronary calcium score (Agatston score) ≥ 1, or CAC-DRS class A1 to A3, is considered indicative of the presence of coronary artery calcification.<br><br>The Agatston score is calculated as the sum of calcified lesion areas within coronary artery projections, multiplied by a density weighting factor based on the peak attenuation (in Hounsfield units, HU) within each lesion:<br><br>Factor 1: 130–199 HU<br><br>Factor 2: 200–299 HU<br><br>Factor 3: 300–399 HU<br><br>Factor 4: ≥400 HU | 0,986 | 0.96 | 0.96 | 0.96 |
| **Epicardial Fat-IRA**<br><br>Assessment of Signs of Ischemic Heart Disease (Epicardial Adipose Tissue) | Automated quantification of epicardial adipose tissue volume on non-contrast chest CT. | Epicardial Adipose Tissue Volume ≥ 125 mL | 0.96 | 1.00 | 0.96 | 0.98 |

| | | | | | | |
|---|---|---|---|---|---|---|
| (ICD-10 Codes I20–I25) | | | | | | |
| **Backbone-IRA**<br><br>AI algorithm for analysis of osteoporosis-related findings (ICD-10 codes M80–M85) | Assessment of vertebral body density to identify imaging features of osteoporosis and osteopenia | Reduced bone mineral density in vertebral bodies from Th11 to L3 (optimally L1–L2).<br>Osteoporosis: < 100 HU<br>Osteopenia: 100–150 HU | 0.995 | 1.00 | 0.98 | 0.99 |
